# Development and evaluation of an interoperable natural language processing system for identifying pneumonia across clinical settings of care

**DOI:** 10.1101/2022.05.10.22274910

**Authors:** Alec B Chapman, Kelly S Peterson, Elizabeth Rutter, McKenna Nevers, Mingyuan Zhang, Jian Ying, Makoto Jones, David Classen, Barbara Jones

## Abstract

**Objective:** To evaluate the feasibility, accuracy, and interoperability of a natural language processing (NLP) system which extracts diagnostic assertions of pneumonia in different clinical notes and institutions.

**Materials and Methods:** An NLP system was designed to identify assertions of pneumonia in three types of clinical notes from electronic health records (EHRs): emergency department notes, radiology reports, and discharge summaries. The lexicon and classification logic were tailored for each note type. The system was first developed and evaluated using annotated notes from the Department of Veterans Affairs. Interoperability was assessed using data from the University of Utah.

**Results:** The NLP system was comprised of 782 rules and achieved moderate-to-high performance in all three note types in VA (precision/recall/f1: emergency=88.1/86.0/87.1; radiology=71.4/96.2/82.0; discharge=88.3/93.0/90.1). When applied to UU data, performance was maintained in emergency and radiology but decreased in discharge summaries (emergency=84.7/94.3/89.3; radiology=79.7/100.0/87.9; discharge=65.5/92.7/76.8). Customization with 34 additional rules increased performance for all note types (emergency=89.3/94.3/91.7; radiology=87.0/100.0/93.1; discharge=75.0/95.1/83.4).

**Conclusion:** NLP can be used to accurately identify the diagnosis of pneumonia in different clinical settings and institutions. A limited amount of customization to account for differences in lexicon, clinical definition of pneumonia, and EHR structure can achieve high accuracy without substantial modification.

## INTRODUCTION

Electronic health records (EHRs) contain detailed information which can be leveraged to improve patient care and support clinical research.[1] However, because the EHR’s primary purpose is to support clinical work, measuring what is meaningful to research and quality improvement is not always straightforward and is impeded by challenges including missing data, inaccurate documentation, and lack of standardization.[2–5] As a result, the information needed to answer many research questions are not found in structured formats and must be looked for in unstructured text.

Natural Language Processing (NLP) can be used to extract relevant data from clinical text. Examples include adverse events,[6–12] social determinants of health,[13–17] and infectious disease surveillance.[18–24] However, a major barrier to using NLP for research is interoperability. Few NLP systems are designed to generalize beyond a particular institution which may have different EHRs or documentation practices.[25,26] Additionally, even within institutions, the vocabulary and structure of a clinical note vary substantially across clinical settings with different types of notes.[27] Thus, many existing tools suffer degraded performance when transported to a new setting, preventing researchers from taking advantage of previously developed NLP tools.

One area ripe for NLP is the diagnosis of pneumonia. Pneumonia is the leading infectious cause of death in the United States.[28–30] Timely and accurate diagnosis and treatment are critical to optimizing prognosis. Incorrect initial diagnosis of pneumonia can lead to poor outcomes including overuse of antibiotics among patients with a false positive diagnosis [31] and delayed treatment among those with a missed diagnosis.[32] Currently, epidemiologic studies of pneumonia must rely on diagnosis codes, which have been shown to have poor accuracy.[33– 37] Previous NLP tools have been developed to extract pneumonia from clinical text, but these largely focused on a single clinical setting or note type and were not scaled across different EHR systems.[38–46]

In this work, we aimed to develop a flexible and generalizable NLP approach for extracting assertions of pneumonia across different clinical settings to improve the measurement of diagnosis. To create a richer and more robust system, we used a diverse sample of annotated notes from the Department of Veterans Affairs (VA) EHR and designed our system to extract assertions of pneumonia from three types of clinical notes: emergency room notes, chest imaging radiology reports, and discharge summaries. To test the generalizability of this method, we then evaluated our system with EHR data from the University of Utah (UU).

## MATERIALS AND METHODS

### Dataset and Participants

We defined our cohort as patients ≥ 18 years who were admitted to the hospital from the emergency department between 1/1/2015-4/30/21 in VA or UU. For the development and initial evaluation of our system, we gathered data from the VA Corporate Data Warehouse (CDW) which is collected from the VA EHR. The VA healthcare system is the largest integrated healthcare system in the United States, with 161 emergency departments and a shared EHR and clinical data warehouse that contains granular clinical data including text from clinical documents and radiology reports that can be queried at the national level. For testing in UU, we retrieved data from the UU Enterprise Data Warehouse (EDW) which is collected from the University’s Epic EHR. The University of Utah consists of a single academic hospital-affiliated emergency department and one additional emergency department with approximately 50,000 patient encounters annually.[47]

The document inclusion criteria and sampling procedure were similar in both systems. Using a combination of keyword searches, document titles, and signing provider specialties, we identified notes for each of the three clinical settings. Keywords and additional details regarding inclusion criteria are included in the Supplemental Materials. Sampled notes were randomly assigned to either a training set, which was used for developing and refining our system, or a testing set, which was used for a final blind evaluation.

### Annotation

Next, we sampled each type of note from VA patient encounters to develop an annotated corpus. Two annotators, an emergency physician (ER) and a pulmonary and critical care physician (BJ), annotated notes and developed consensus annotation guidelines. For each note type, the annotation process was divided into two phases. First, annotators iteratively reviewed identical batches of 10-20 notes each, measured inter-annotator agreement (IAA), and resolved disagreements. Once an acceptable IAA had been reached, defined as Cohen’s Kappa >= 0.8,[48] the annotators proceeded to review notes separately using the agreed upon guidelines. For each note type, 100 documents were annotated by both reviewers and used as the testing set for NLP development. Final IAA was measured using this double-annotated VA testing set.

Annotators were instructed to classify each document as either “Positive”, which included both definite and possible diagnoses of pneumonia, or “Negative”. Definitions of a pneumonia diagnosis varied slightly between each of the three note types, accounting for the different document structures and varying levels of diagnostic uncertainty at each step of the hospitalization. A summary of classification logic for each document type is given below. Additionally, the complete annotation guidelines are included in the Supplemental Materials.

#### Emergency Department

A document was classified as “Positive” if the author documented an assertion of pneumonia in a relevant section without ruling it out using radiographic findings. Annotators first searched the “Assessment/Plan”, “Diagnosis”, or “Addendum” sections for a final diagnosis. If there were no diagnostic assertions in any of these sections, annotators next reviewed the “Medical Decision Making” and “ED Course” for relevant information.

#### Radiology Report

A report was classified as “Positive” if there was an assertion of pneumonia or radiographic findings which were supportive of pneumonia. These included clinical terms for pneumonia as well as radiographic terms including “infiltrate”, “airspace disease”, “consolidation”, “opacity”, and “interstitial process”. Some radiographic findings could be listed as evidence of other diagnoses such as atelectasis or interstitial lung disease. In these cases, annotators classified the document as “Negative”.

#### Discharge Summary

A document was classified as “Positive” if there was an assertion of pneumonia in either the “Diagnoses” or “Hospital Course” sections. Particularly in the hospital course, a distinction had to be made between initial and final diagnoses of pneumonia. For example, a document containing the text “<u>Upon admission</u> the patient was diagnosed with **pneumonia** but <u>CHF appears more likely</u>” would be annotated as “Negative”.

### NLP System

We implemented our system using the Python package medspaCy which extends the popular platform spaCy^1^ for clinical text processing.[49] We chose to design our system as a rule-based pipeline due to the highly specific criteria needed to classify notes in each domain, as well and the relatively small amount of annotated data. Our code is publicly available on GitHub.^2^ An example of a discharge summary processed by our system is shown in **Figure 1**.

**Figure 1.**
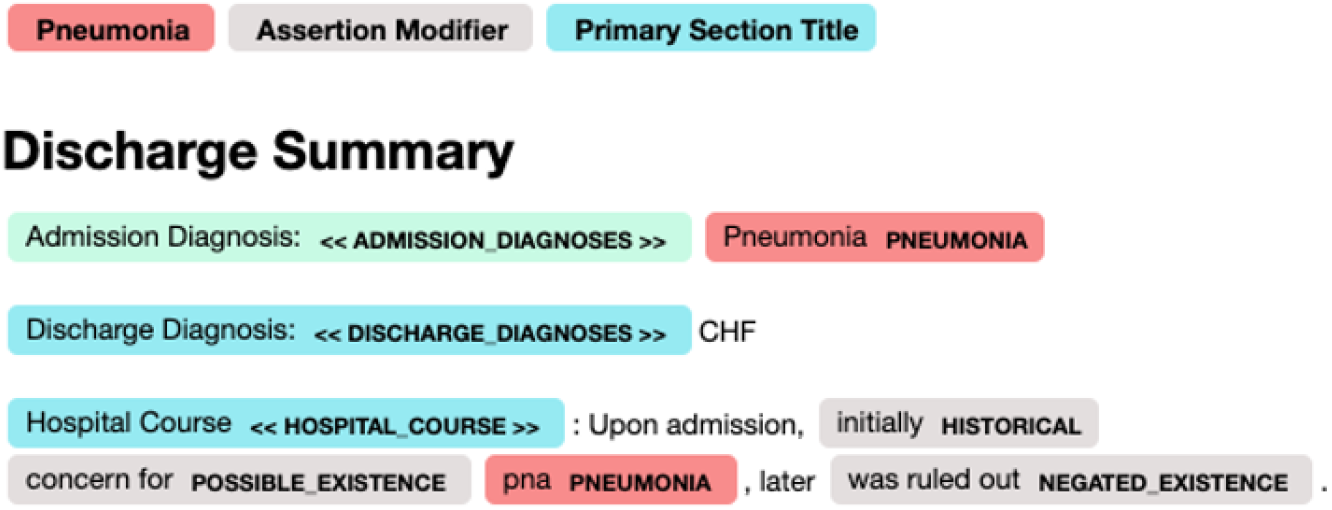
Visualization of a discharge summary processed by the NLP.

### Flexible System Architecture

To account for the variation in lexical content and clinical definitions, we aimed to design an NLP system which could be modified as needed for specific note types and different institutions while keeping the primary logic consistent across all settings. The modular and customizable structure of medspaCy pipeline components allowed us to organize our model accordingly.

Our system allowed for this flexibility by organizing separate knowledge bases containing either “common” or setting-specific resources. Common resources defined general lexical terms, section titles, and linguistic patterns, while setting-specific resources implemented rules for each particular note type and institution. The overall design of our pipeline is shown in **Figure 2**.

**Figure 2.**
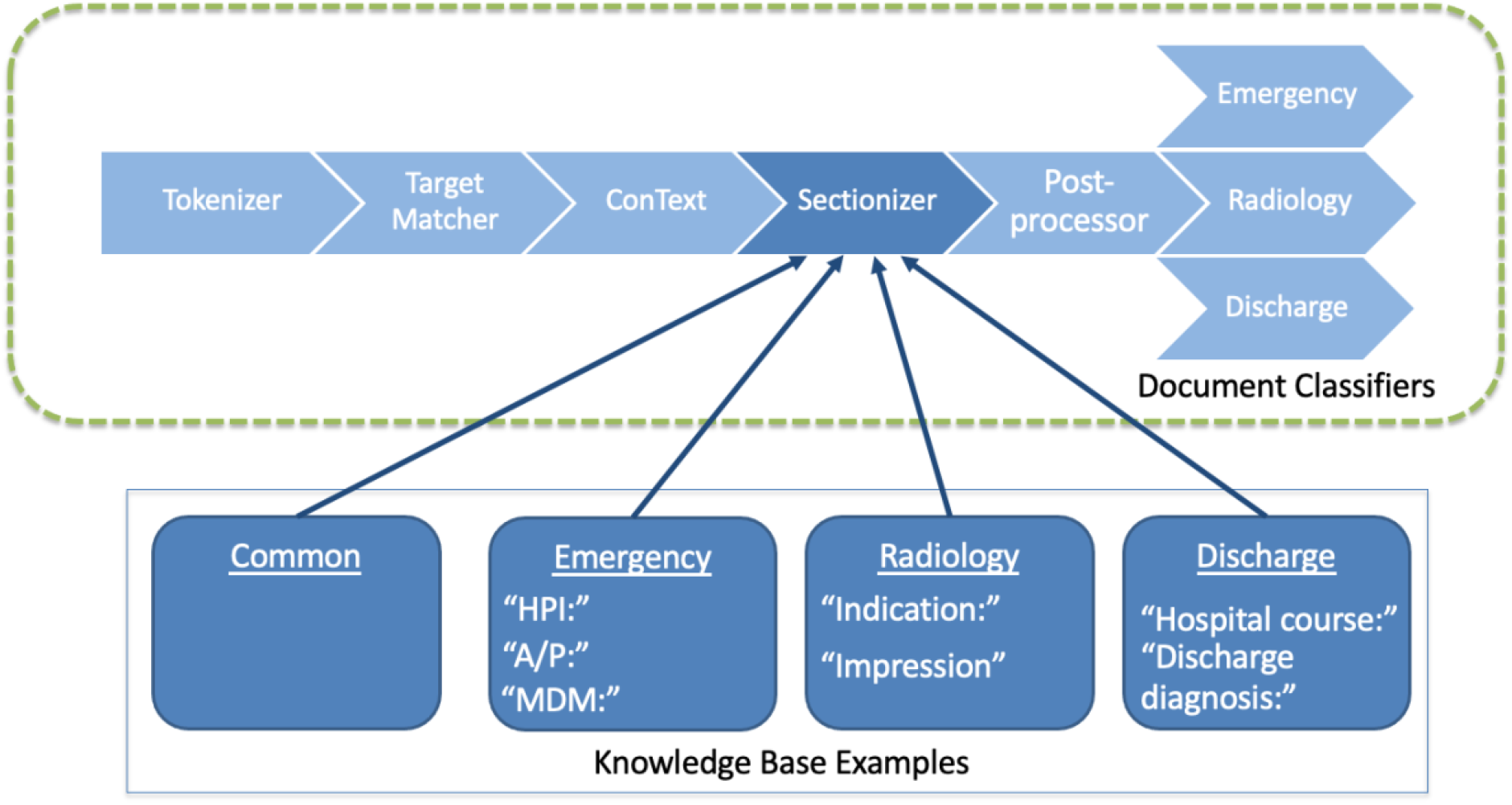
Conceptual representation of the modular NLP system design.

One purpose of customization was to allow for prioritization of different performance characteristics for each clinical setting to match the intended purpose for each note type. For example, since the standard of care for pneumonia is to confirm a clinical diagnosis by chest imaging, we felt that the primary purpose of a radiology report is to confirm positive clinical diagnoses of pneumonia.[50] As such, we tailored our classification system to prioritize recall over precision. By contrast, we planned on using ED and DC notes for identifying both positive and negative diagnoses and prioritized a balance between precision and recall for those notes.

### Entity Extraction

Following initial standard processing steps such as tokenization, part-of-speech tagging, and dependency parsing, the first step in our system was entity extraction using medspaCy’s *TargetMatcher* component. Spans of text representing pneumonia and related concepts were identified in the text using semantic and syntactic patterns. Examples of concept labels included “pneumonia”, “opacification”, “anatomy”. A number of rules were adapted from previous work using the NLP system Moonstone.[15]

Some examples of setting-specific entity rules were “airspace disease” and “infectious process”. These phrases were included in the RAD pipeline because they have specific meaning in this setting but were excluded from the ED and DC pipelines as they were ambiguous in clinical notes. Additionally, the RAD pipeline extracted concepts pertaining to alternate diagnoses such as atelectasis and pulmonary edema, while phrases like “Klebsiella pneumoniae” and “pneumonia vaccine” needed to be tagged as separate entities in ED and DC but were not needed in RAD.

### Attribute Detection

The ConText algorithm was used to identify attributes including negation, uncertainty, temporality, and hypothetical assertion from neighboring text.[51] An example of how ConText links entities to relevant linguistic modifiers is shown in **Figure 3**.

**Figure 3.**
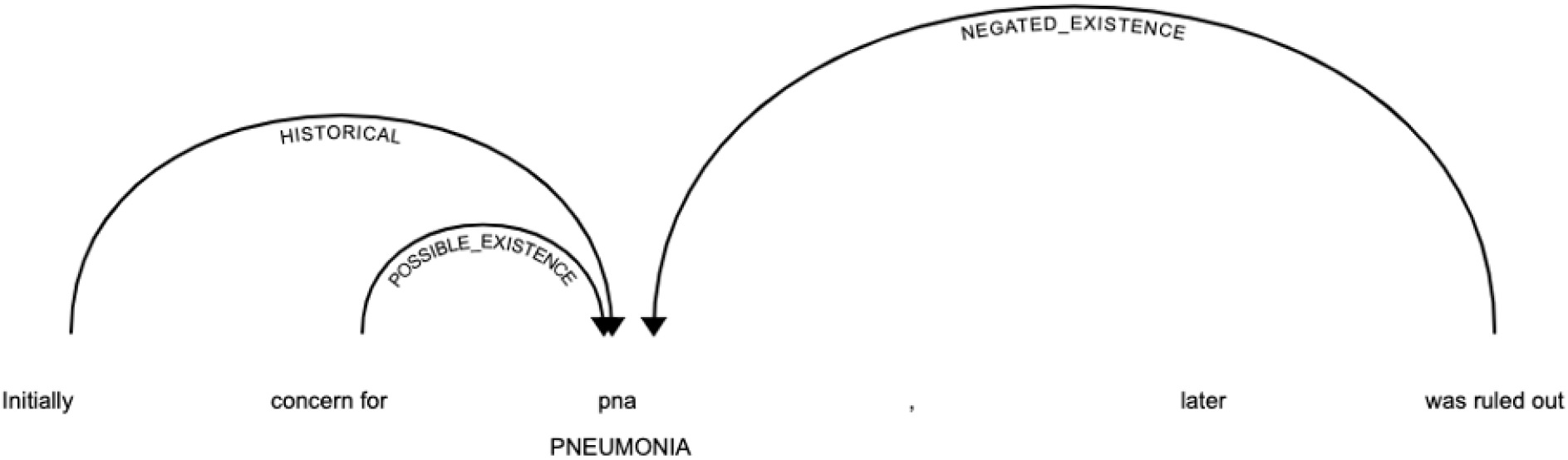
The ConText algorithm detecting linguistic attributes for a mention of pneumonia.

Some phrases which may be used to assert a possible diagnosis in the clinical pipelines needed to be handled differently in the RAD pipeline. For example, in an ED note, the phrase “rule out pneumonia” would indicate that the author suspects pneumonia may be a diagnosis and thus would be classified as “Positive”. However, in a chest imaging report, the phrase “rule out” pneumonia would only give an indication for the reason for the procedure and would not relay anything about the radiologist’s interpretation of the findings. In this context, “rule out” was treated as hypothetical and would not merit a positive classification.

Similarly, in DC summaries, the phrase “initial” typically referred to the diagnosis at time of admission rather than the final discharge diagnosis. Accordingly, a rule matching this phrase was included in the DC pipeline but not the ED pipeline.

### Section Detection

Since the annotation guidelines explicitly refer to different sections of a note for each domain, identifying which section of a note contained a mention of pneumonia was crucial for correctly classifying each note. Because the VA EHR does not enforce standardized note templates, there were many different variations of section titles which needed to be captured in pipeline. The need for detecting section titles was particularly important for emergency notes and discharge summaries. We used medspaCy’s *Sectionizer* component to identify sections.

### Postprocessing

The medspaCy *Postprocessor* component executes custom business logic on the entities in a document. One important postprocessing rule was disambiguating “CAP” in the clinical notes as meaning either “community-acquired pneumonia” or referring to a form of medication. Another postprocessing rule ignored irrelevant mentions of opacification in radiology reports by linking them to an anatomical site of the pulmonary artery (e.g., “suboptimal ***opacification*** of the <u>pulmonary artery</u>”).

### Document Classification Logic

A custom document classifier component was implemented separately for each domain. Each document classifier attempted to replicate the logic specified in the annotation guidelines based on the entities and corresponding attributes in the document. The primary difference between classification logic for ED and DC notes was which sections were considered for a “Positive” classification. The radiology system included radiographic concepts in classification logic and attempted to exclude findings which were supporting an alternate diagnosis such as atelectasis or pulmonary fibrosis.

### Customization for Generalizability

VA and UU use different EHRs (VistA and Epic, respectively) and have different note structures. Additionally, while VA has multiple sites across the country, each with the ability to customize and modify note structure, UU consists of a single site and generally has less variation in formatting. It was therefore necessary to modify the NLP system to match the UU note structure. However, we hypothesized that the general logic in each institution would not need to be modified and that a small amount of customization in the NLP design could effectively adapt to the new institution. To implement these customizations, we created a separate set of knowledge base files for UU data. Most additional resources were additional section headers which matched the structure of Epic notes, as well as some rules to match entity terms or modifiers which were not seen in VA notes. We did not modify the document classification logic.

### Evaluation

Following development of our NLP system, we next evaluated its performance at identifying pneumonia in each note type. We performed a total of three evaluations. First, we measured performance using the VA testing set. Next, we performed two evaluations using the UU testing set. We first ran the model “out-of-box” with no modifications to test how well our system could generalize to a new system without additional development effort. Finally, we evaluated a customized system including UU-specific resources which had been developed using a small set of training documents. For each evaluation, we measured binary classification performance using precision, recall and F1^3^.

### Error Analysis

Among all testing documents with an error in any of the three evaluations, we conducted an error analysis. Each case was reviewed by an NLP developer (AC) to identify sources of NLP error, and cases with ambiguity were additionally reviewed by a clinician (BJ). We identified general categories of errors and tabulated the total number of errors by category in each evaluation.

## RESULTS

### Dataset and Patient Cohort

Summary statistics for counts of patients in our cohort and relevant notes are displayed in

### Annotation

For the VA training set, annotators annotated a total of 377 ED notes, 267 RAD reports, and 200 DC summaries. For the VA testing set, 100 notes of each type were double annotated with disagreements reviewed and resolved by consensus. The final agreement on the testing set was Cohen’s Kappa = 89.9 for ED, 94.9 for RAD, and 83.9 for DC.

For the UU dataset, one annotator annotated 100 documents of each note type for training while the other annotator annotated 100 documents of each for testing. Notes were not double annotated; instead, a small batch of 40 practice documents from each domain were annotated beforehand with a high enough agreement for each note type (Kappa > 0.8) to establish confidence in annotator reliability using the UU dataset.

### NLP Evaluation

The results for the NLP system on the VA testing set are shown in **Table 2**. Our system achieved high precision in ED and DC, moderate precision in RAD, and high recall in all three domains.

**Table 1.** Summary statistics for our dataset and cohort in both institutions.

|  | VA | UU |
| --- | --- | --- |
| Count of patient encounters in cohort | 2,203,165 | 81,144 |
| Date range of notes | 1/1/2015-4/30/2021 | 1/1/2015-4/30/2021 |
| Total count of notes with keywords |  |  |
| Emergency | 1,219,868 | 18,931 |
| Radiology | 1,846,246 | 82,051 |
| Discharge | 2,130,274 | 10,506 |

**Table 2.** NLP performance on the VA testing set (n=100 documents per note type).

| VA |  |  |  |
| --- | --- | --- | --- |
|  | Precision | Recall | F1 |
| Emergency | 88.1 | 86.0 | 87.1 |
| Radiology | 71.4 | **96.2** | 82.0 |
| Discharge | **88.3** | 93.0 | **90.1** |
| Macro Avg | 83.9 | 91.2 | 87.4 |

The highest overall F1 score was achieved in DC. Precision and recall were fairly balanced in both ED and DC. Radiology reports were less precise but more sensitive. These relative performance characteristics matched our objectives for prioritizing precision or recall in each setting. A total of 782 rules were included in our VA pipeline (307 target concept rules, 288 section rules, 178 context rules, and 9 postprocessing rules).

The performance of our system using UU data is shown in **Table 3**. Testing the NLP “out-of-box” yielded overall comparable performance to VA. Recall was quite high in all domains, increasing for both ED and RAD and dropping only slightly for DC. Precision increased in RAD and saw a small decrease for ED. The largest drop in performance was precision for DC. Customizing the NLP for UU data improved performance for all note types and metrics and achieved generally higher performance than in VA. The only metrics which were lower than in VA were precision and F1 for DC. The customized UU system included an additional 34 rules.

**Table 3.**
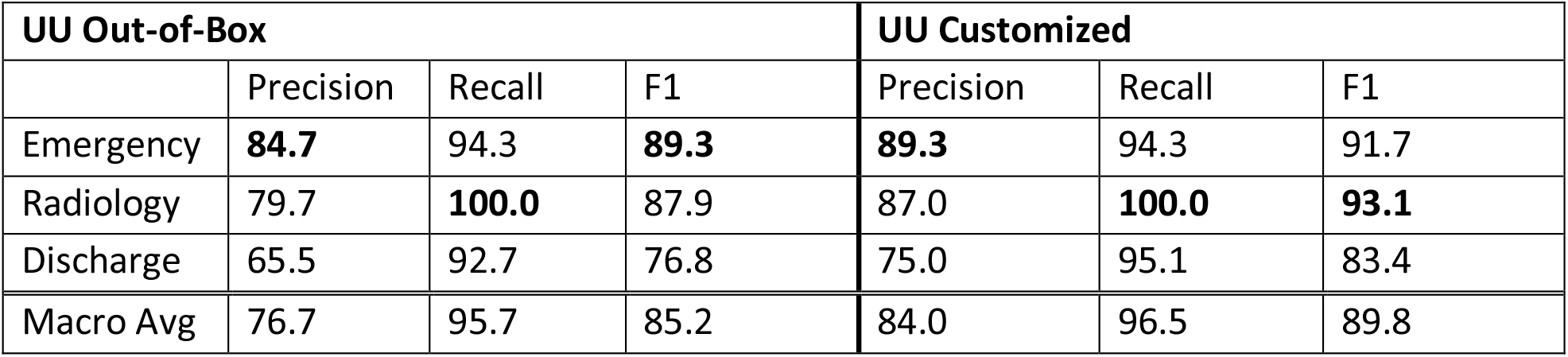
NLP performance using UU data (n=100 documents per note type). “UU Out-of-Box” performance using the exact system developed in VA, while “UU Customized” was customized for UU data.

| UU Out-of-Box |  |  |  | UU Customized |  |  |
| --- | --- | --- | --- | --- | --- | --- |
|  | Precision | Recall | F1 | Precision | Recall | F1 |
| Emergency | **84.7** | 94.3 | **89.3** | **89.3** | 94.3 | 91.7 |
| Radiology | 79.7 | **100.0** | 87.9 | 87.0 | **100.0** | **93.1** |
| Discharge | 65.5 | 92.7 | 76.8 | 75.0 | 95.1 | 83.4 |
| Macro Avg | 76.7 | 95.7 | 85.2 | 84.0 | 96.5 | 89.8 |

### Error Analysis

Within the 300 testing documents, we identified a total of 111 NLP errors, including 33 VA, 47 out-of-box UU, and 31 customized UU. The counts in each domain or error and institution are shown in **Table 4**.

**Table 4.**
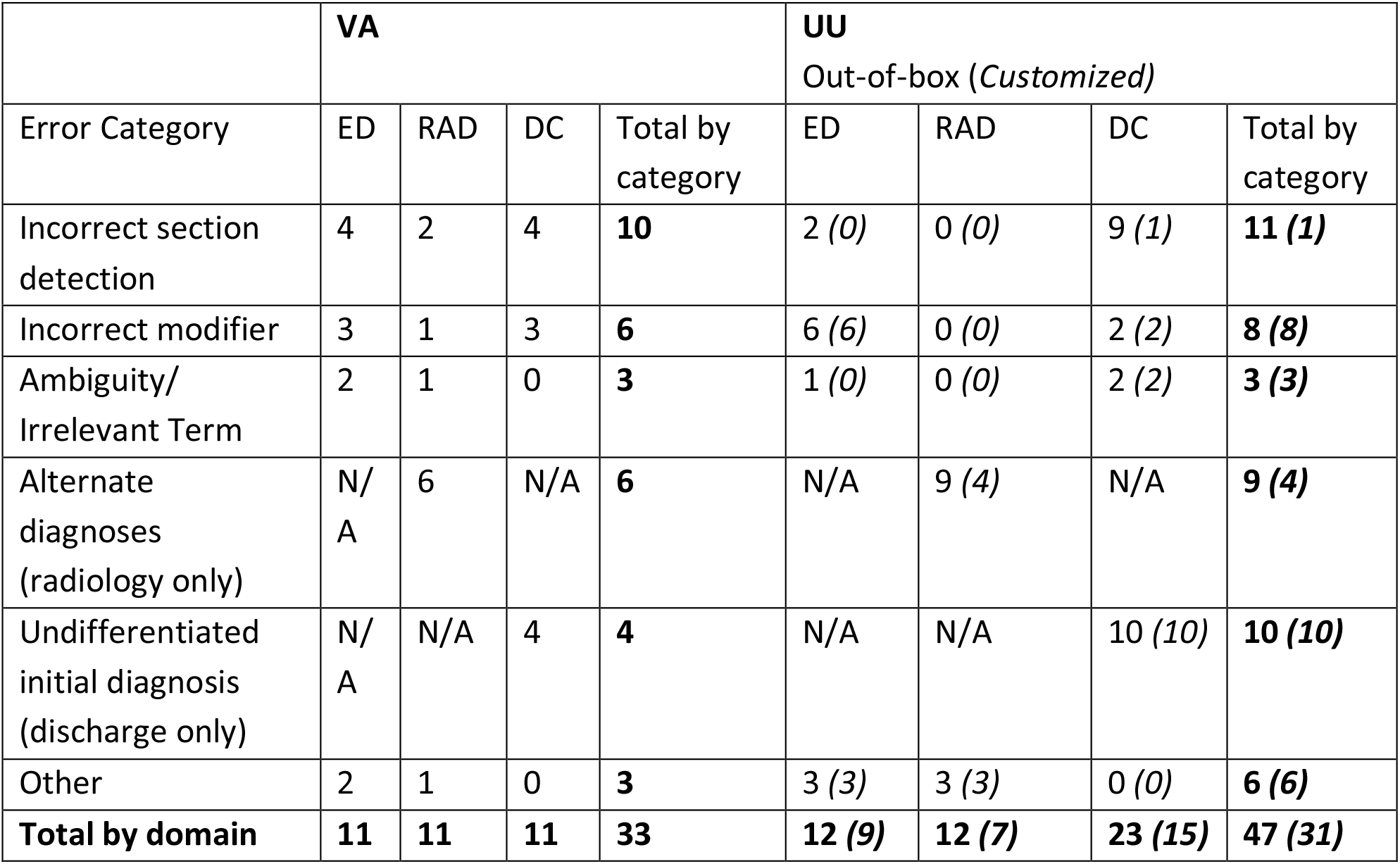
Types of NLP errors in VA and UU testing sets. Counts for UU after customization are shown in *italics*.

| Error Category | VA |  |  |  | UU<br>Out-of-box (*Customized*) |  |  |  |
| --- | --- | --- | --- | --- | --- | --- | --- | --- |
|  | ED | RAD | DC | Total by category | ED | RAD | DC | Total by category |
| Incorrect section detection | 4 | 2 | 4 | **10** | 2 (*0*) | 0 (*0*) | 9 (*1*) | **11 (*1*)** |
| Incorrect modifier | 3 | 1 | 3 | **6** | 6 (*6*) | 0 (*0*) | 2 (*2*) | **8 (*8*)** |
| Ambiguity/<br>Irrelevant Term | 2 | 1 | 0 | **3** | 1 (*0*) | 0 (*0*) | 2 (*2*) | **3 (*3*)** |
| Alternate diagnoses<br>(radiology only) | N/A | 6 | N/A | **6** | N/A | 9 (*4*) | N/A | **9 (*4*)** |
| Undifferentiated initial diagnosis<br>(discharge only) | N/A | N/A | 4 | **4** | N/A | N/A | 10 (*10*) | **10 (*10*)** |
| Other | 2 | 1 | 0 | **3** | 3 (*3*) | 3 (*3*) | 0 (*0*) | **6 (*6*)** |
| **Total by domain** | **11** | **11** | **11** | **33** | **12 (*9*)** | **12 (*7*)** | **23 (*15*)** | **47 (*31*)** |

Across all note types, the most common error in both institutions was incorrect section detection, where a section title was either missed due to non-standard section titles or assigned the wrong classification. This was most common in clinical notes and was especially prevalent when running the “out-of-box” system in UU. For example, UU DC notes frequently had a section titled “Patient Active Problem List Diagnosis” which represented the “Problem List”. However, because this formatting was not present in VA, this section was by default tagged as “Diagnoses” and led to false positives prior to customization.

While error was low for radiology reports, the most common error for this note type was failure to link radiographic findings to an alternate diagnosis, leading to false positives. Some of these instances were fixed in the custom implementation after identifying additional phrases used to link findings to alternate diagnoses.

For DC summaries in UU, a common source of error for both the “out-of-box” and customized systems was confusion between initial and final diagnoses. Most of these cases were false positives extracted from the “Hospital Course” which were documenting earlier diagnostic possibilities but did not reflect the final diagnosis. This error also occurred in VA but was much less common. The “Hospital Course” section in UU tended to be longer and more detailed, often containing much more background information about a patient’s condition and a more detailed narrative about their hospitalization, making it difficult for the NLP to differentiate between the author’s final diagnostic assertion and intermediate diagnoses during the hospitalization.

Other issues seen in each of the notes included incorrect modifier linking, ambiguous terms, patient instructional text, and some cases of annotation error.

## DISCUSSION

### Key Findings

Using the clinical text from two healthcare and EHR systems, we developed and tested a flexible, generalizable Natural Language Processing system that can accurately identify diagnostic assertions of pneumonia across different clinical settings and institutions. We found that some customization to accommodate formatting differences in a new institution improved performance. However, high performance was achievable by adding only a small number of rules. Performance was particularly high for chest imaging and emergency department notes, even without modification, suggesting that interoperable NLP tools for identifying pneumonia within multiple institutions are feasible.

### Related Work

Our work builds upon existing studies of NLP in pneumonia but represents several important advances. First, the majority of previous work has focused on extracting radiographic evidence of pneumonia from radiology reports, [38–40,42,43,45] which does not shed much light on clinical diagnoses. Similar to our radiology pipeline, most of these studies achieved moderate precision and higher recall. There are far fewer studies utilizing text from care teams to identify clinical diagnoses. Bejan et al developed a machine learning algorithm to identify pneumonia diagnoses among intensive care unit patients using features extracted from multiple note types and found moderate accuracy at the patient level.[46] Their study focused on a small sample of patients from a single institution. Jones et al used a combination of structured data and a machine-learning NLP from the VA system to classify initial diagnoses of pneumonia from emergency department notes,[41] finding that NLP achieved much higher recall than ICD codes and that combining NLP with ICD codes, which had high precision, achieved the highest overall score. An important finding of this study was that NLP identified more uncertain cases of pneumonia, which is important when studying the evolution or change in diagnosis for an individual patient across an episode of care. None of the previous studies tested the NLP system outside of the institution in which it was developed.

To our knowledge, our study is the first to assess the generalizability of pneumonia NLP across different clinical settings, institutions, and EHR systems. Generalizability is important, particularly when developing measures that are used to examine variation of phenomena at a large scale. Currently, much of measurement in healthcare relies upon chart review, which is extremely burdensome and variable and has led to a call for electronic clinical quality measures (eCQMs).[52] In order for eCQM’s to be useful, they must represent the same constructs of quality across institutions.

Our system uses rule-based rather than machine learning methods, while other studies have demonstrated promise in machine learning.[39–43,45,46] We chose a rule-based design for this project for several reasons. Surveys comparing rule-based to machine learning approaches have shown several practical advantages to rule-based NLP, including interpretability, the ability to incorporate domain knowledge, the relatively simple process of modifying rules for new use cases and datasets, and the smaller amount of labeled data required for training.[53] The modifiable nature of rules also allowed us to easily update the system to fit new data, allowing us to customize our system to different note types and institutions. Perhaps the most important advantage, however, was the usability of rule-based NLP to human learning. An important purpose of our system is to provide measures that support clinician review of cases in which diagnoses change across a hospitalization in order to identify opportunities to improve their diagnostic skills. Using an approach that is interpretable, explainable, and scrutable helps users to understand the underlying logic of the system,[54,55] which is essential for building transparency and trust among clinicians during feedback or decision support.[56] Combining explainable machine learning with rule-based approaches to generate measures that are simultaneously highly accurate, resilient to changes in data, and interpretable, as well as meaningful across multiple institutions, is the subject of future work.

The findings in this study have several important implications on research and quality improvement aimed at improving diagnosis within a learning healthcare system. The accuracy and generalizability of the proposed system demonstrate the potential of using NLP to measure the evolution of diagnosis across clinical settings and within multiple institutions, which can be used at the system and provider level for learning. As the EHR becomes increasingly ubiquitous and powerful in clinical care, it is essential that we leverage it to support learning. Since a large amount of EHR data is recorded in clinical text, important elements that are not easily coded, such as diagnosis, can be lost unless we develop automated approaches through NLP. If we are to truly leverage the clinical experience from millions of patient encounters, we must develop NLP methodologies that are simultaneously accurate, interpretable to humans, and designed for meaningful improvement.

### Limitations

We recognize several opportunities to improve of our system that were not tested in this study. While the performance of our NLP system was generally high, continuing to refine section detection – either through additional rules or through the development and use of more standardized note template – and modifier linking are important ways to further improve accuracy. Combining this text processing system with structured data such as diagnosis codes and medical treatment could increase positive predictive value of pneumonia identification, particularly for discharge diagnoses in UU data. While our study used three note types to capture diagnoses in three distinct settings across a patient’s hospitalization, there are additional types of notes such as progress notes, admission history and physicals, consultation reports, even follow-up outpatient notes that may further enhance our understanding of the evolution of diagnosis across a patient’s care episode. Since the system can accommodate additional note types, future work includes expanding this approach to include new note types. Similarly, while the scope of this study was limited to the diagnosis of pneumonia, future work can extend this approach to other diseases, particularly those that can mimic pneumonia such as heart failure.

## CONCLUSION

Using clinical text from two separate institutions and EHRs, we demonstrated the feasibility, accuracy, and generalizability of a flexible NLP system that extracts assertions of pneumonia diagnoses across different clinical settings, healthcare institutions, and EHR systems. We are currently applying this system to examine the process of pneumonia diagnosis across hospitalizations and develop measures and tools that support provider and system-level feedback and learning. By unlocking data from clinical text, NLP has enhanced our ability to characterize the process of diagnosis at a large scale. This capability will advance our understanding of the process of diagnosis in healthcare and how to best support diagnostic excellence.

## Data Availability

Data are not available per IRB protocol.
Code is publicly available on GitHub.

https://github.com/abchapman93/medspacy_pneumonia

## ACKNOWLEDGEMENTS

The authors thank Battelle Memorial Institute, Julie Varner, Vikrant Deshmukh, Lee Christensen, Dr. Wendy Chapman, Patrick Alba for their support and contributions to this work.

## AUTHOR CONTRIBUTIONS

AC and KP contributed to study design, NLP system development, data curation, data analysis, system evaluation, and manuscript preparation. ER contributed to conceptualization, study design, annotation, and manuscript preparation. MN contributed to data curation, data analysis, and manuscript preparation. MZ contributed to data curation and manuscript preparation. JY contributed to study design, data analysis, and manuscript preparation. MJ and DC contributed to conceptualization, study design, and manuscript preparation. BJ contributed to conceptualization, study design, data curation, data analysis, annotation, system evaluation, and manuscript preparation.

## Footnotes

1 https://spacy.io/

2 https://github.com/abchapman93/medspacy_pneumonia

3 Precision = (# TP) / (# TP + # FP); Recall = (# TP) / (# TP + # FN); F1 = 2*Precision*Recall / (Precision + Recall) TP = true positives; FP = false positives; FN = false negatives

