## Supplemental Materials for "Development and evaluation of an interoperable natural language processing system for identifying pneumonia across clinical settings of care"

**Table of Contents**

I. Code

II. Document Inclusion Criteria

III. Emergency Note Annotation Guidelines

IV. Radiology Report Annotation Guidelines

V. Discharge Summary Annotation Guidelines

I. Code

**Link to code:** <https://github.com/abchapman93/medspacy_pneumonia>

We implemented our system in Python using [medspaCy](https://github.com/medspacy/medspacy). A version of the code, including all knowledge base files and rules for both VA and UU, can be found in the GitHub repository. Our code can also be installed as a Python package using pip:

pip install medspacy_pna

You will also need to install spaCy’s en_core_web_sm model to use the default pipeline settings:

python -m spacy download en_core_web_sm

This package requires Python 3.8, medspaCy 0.2.0.0, and spaCy 3.1.*.

II. Document Inclusion Criteria

Emergency Department (ED) notes were authored by an emergency medical doctor, nurse practitioner, or physician assistant (MD/DO, NP or PA) and had a title such as “Emergency Department Note”. To identify notes relevant to our task, we searched for clinical pneumonia terms such as “pneumonia”, “pna”, and other variations. In particular, we did not include radiographic terms in our ED keyword search as those terms were found to return many notes with negative chest imaging findings but no clinical suspicion for pneumonia. Finally, we identified any addenda and concatenated them to the original ED note, as these addenda often contained the physician’s final assessment of the patient.

For RAD reports, we extracted chest x-ray (CXR) and computed tomography (CT) reports. The keyword search for RAD reports included clinical terms for pneumonia as well as related radiographic terms such as “infiltrate” and “consolidation”.

For discharge summaries (DC), we included notes which were authored by a MD/DO, NP or PA and had a title such as “Discharge Summary” while excluding notes with “Instructions” or “Education” in the title. We filtered out patient education notes and discharge instructions. We used the same clinical keywords for DC summaries as we had for ED notes.

**Keywords**

| All note types | RAD only |
| --- | --- |
| Pneumonia  Pneumonie  Pneumonias  Pna  Hcap  Bkp  Bronchopneumonia  Pleuropneumonia | Consolidation  Infiltrate  Opacification  Consolidate  Opacity  Opacities  Opacified  Pneumonitis  Airspace disease  Air space disease |

III. Emergency Note Annotation Guidelines

Thank you for helping to annotate for this project! Our goal is to develop a natural language processing tool that can classify clinician-written documents on whether they contain a positive assertion of the diagnosis of pneumonia.

### 1. **Background**

**The initial emergency department note** is a clinical document written by physicians, nurse practitioners, or physician assistants who are responsible for diagnosing pneumonia in the emergency department

Other document types that may eventually be tested include:

1. Admission History & Physical (written by admitting hospital care team)
2. Progress note (daily notes written by hospital care teams or consultants)
3. Consultation Report (initial note written by consulting service at the time of consultation)

**The question we are asking is:**

***“Does the clinician/author of the note states/assert that the patient has a positive or possible diagnosis of pneumonia?***

We are not asking:

- “Does the patient actually have evidence for pneumonia?” / “Do *you* think the patient actually has pneumonia?”
- “Does the patient have a history of pneumonia?”
- “Did the provider *treat* the patient as though the patient had pneumonia (ie did not mention pneumonia, but mentioned respiratory antibiotics)”?
- “Did the clinician clearly rule out or exclude pneumonia?” - The task of this classifier is to identify whether the clinician asserted pneumonia as a leading or possible diagnosis within the assessment and plan.” We are looking for *positive or* *possible* assertions of a pneumonia diagnosis. The task is NOT to accurately identify clearly negative assertions. (ie in our project, the “negative” category just means lack of assertion of a pneumonia diagnosis, rather than an assertion that pneumonia was ruled out or excluded.)

### 2. **General Guidelines**

#### **2.1 Definitions: Positive, possible, or negative**

1. POSITIVE = Clinician states with certainty of the diagnosis of pneumonia without mentioning any other possibilities.
2. POSSIBLE = Possible pneumonia– clinician states the dx of pneumonia is possible or probable but uses terms of uncertainty or lists other diagnoses as alternatives.
3. NEGATIVE = Clinician indicates a different diagnosis and does not list pneumonia as a possible diagnosis, or mentions that pneumonia was specifically excluded. Note: in this case, we are treating absence of evidence as evidence of absence… ie, “negative” just means a lack of positive/possible classification. (see below in the “inconsistencies/ambiguities” section for more details).

#### **2.2 Summary of classification logic**

- A document is classified as **“Positive”** if pneumonia is definitively asserted as being the clinical diagnosis within a “Tier 1” section such as “Assessment/Plan” or “Final Diagnoses”
- A document is classified as **“Possible”** if:
  - Pneumonia is listed as a possible diagnosis in a “Tier 1” section
  - **Or** pneumonia is listed as a diagnosis (either definitively or with uncertainty) in a “Tier 2” section such as “Medical decision making” or “Differential diagnosis” **and** is not ruled out elsewhere in the document
- A document is classified as **“Negative”**
  - Pneumonia is not listed as a diagnosis in a “Tier 1” or “Tier 2” section
  - **Or** pneumonia is listed as a possible diagnosis in a “Tier 2” section but is later ruled out either explicitly (e.g., “pneumonia is ruled out”) or implicitly by radiographic findings (e.g., “Chest x-ray did not show any infiltrate or consolidation”)

This logic is shown visually below. See **Section 3** for detailed terms and sections.

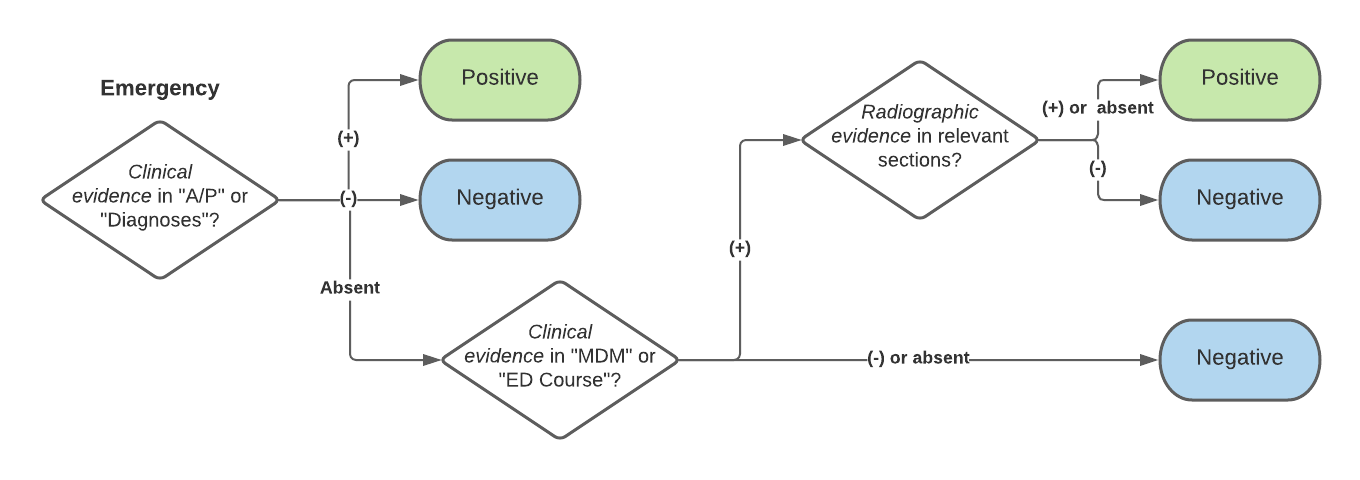

#### **2.3 Suggested workflow**

1. Find all relevant A/P sections (Assessment/Plan, Plan, Impression, Diagnosis, Medical Decision-Making/differential diagnosis, ED course- see Table 1 for details).
2. Within these sections, search for clinical pneumonia terms.
3. If clinical pneumonia is mentioned in any section and not clearly excluded by a subsequent section, classify positive or possible accordingly.

#### **2.4 General principles**

- Use clinician’s narrative (assertion/suspicion within an assessment/plan), rather than objective information (evidence).
- Location matters: the place in which the relevant information depends upon the note type.
- Use what clinicians say, not what you think they mean. Ie, the task is to classify the clinicians’ statement/assertions of the diagnosis, but avoid inferring the diagnosis from their treatment decisions (ie, order CXR, treat with antibiotics).
- Negative radiographic findings may be used to rule out a possible diagnosis of pneumonia **if** the radiographic findings occur in a relevant section (e.g.,embedded in the A/P rather than a separate imaging section).
- Positive radiographic findings may not be used to classify a document (e.g., “Chest x-ray showed infiltrate” does not qualify as positive)
- We are not trying to distinguish between new and existing cases of pneumonia. So a diagnosis of healthcare-associated pneumonia acquired in a previous hospitalization could still be considered “Positive”

### 3. **Specific terms and sections**

#### **3.1 Terms used for pneumonia**

| Clinical terms of pneumonia  *These may be used by clinician or radiologist | Pneumonia and misspellings  HCAP  PNA  Bkp  bronchopneumonia  cap-  legionellosis  Parapneumonia effusion  Empyema  Pneumonia protocol |
| --- | --- |
| Radiographic terms of pneumonia  *These are more often used by radiologist, are non-specific to pneumonia; are positive for chest imaging, but not necessarily for ED clinician note. | Opacity  Infiltrate  Consolidation |
| Terms that may imply pneumonia but should not be used as specific pneumonia terms | Atypical infection  Respiratory infection  LRTI (lower RT infection)  Lung infection  Sepsis, lung source  Community-acquired |

Parapneumonic effusion = collection of fluid between lung and chest wall (the pleural space). Empyema = collection of pus between lung and chest wall.

If a clinician includes “infection/empyema” or “parapneumonic effusion” in their diagnosis, they are stating the patient has pneumonia.

#### **3.2 Linguistic modifiers**

Linguistic modifiers are phrases that refer to a mention of pneumonia and express uncertainty, negation, or temporality. When reviewing a mention of pneumonia, you should review for any of these terms to see if the mention of pneumonia should be excluded.

| **Modifier category** | **Examples** |
| --- | --- |
| Positive existence | positive for  resolving  Improving  Consistent with  Likely  Suspicious for |
| Possible existence | differential diagnosis  possible  Pneumonia vs *X*  rule out  less likely |
| Negative existence | ruled out  no signs of  risk of pneumonia is low  is not likely  doubt  Excluded  Does not clinically correlate  Low suspicion for  Low concern for |
| Historical (annotate as “Negative”) | recent  past medical history of  resolved |

#### **3.3 Relevant sections**

Sections are divided into two levels:

- **Tier 1:** Sections containing final clinical and diagnostic decisions. Tier 1 sections include the Assessment/Plan and final diagnoses and are often near the end of a note. If there is information regarding pneumonia in a Tier 1 section, this will typically supercede any information in a Tier 2 section.
- **Tier 2:** Sections containing intermediate decision making or information, such as “Medical decision making” or “Hospital course”

Note: these sections are often interchangeable, sometimes with one or more missing. They typically are sequenced in an order that follows the clinical workup, so that a diagnosis that is considered in earlier sections might be ruled out/excluded in a later section. **If any assertion is made of pneumonia within these sections, this should be classified as positive or possible *unless following sections clearly state that the diagnosis was excluded.***

***Examples of exclusions are:***

- **Clear descriptions of negating pneumonia**
- **Including the chest imaging report within the relevant section that negates pneumonia using clinical or radiographic terms (“negative for pneumonia,” “no consolidation”)**

Sections NOT to use when classifying:

- Do not use chief complaint or reported symptoms
- Do not use chest imaging report results
- Do not use physical exam findings
- Do not use vitals/labs.
- Do not use medication lists that are not in the plan (ie the medications the patient was taking prior to the visit)
- Do not use patient instructions/hypothetical (return to ER if…)

If the note is clearly not a physician note (ex, it is an addendum showing XR result, or nursing not, or lab results etc), classify as “Not MD note” and do NOT annotate.

Relevant Tier 1 and Tier 2 tables are summarized in the table below.

| **Tier 1** | Assessment/Plan  Diagnoses/Final Diagnoses  Impression (*not* imaging report impression)  Addendum |
| --- | --- |
| **Tier 2** | Medical decision making  Differential diagnoses |

#### **3.4 Inconsistencies/ambiguities**

**a. Evolving or differing diagnoses in the MDM and A/P sections:** Often, the diagnosis evolves during the ED course and this is reflected in the process of writing the note. Emergency department note can contain a “medical decision-making/emergency department course” section, which lists an *initial* list of possible or differential diagnosis before all of the studies/work-up comes back. This working diagnosis can be inconsistent with the clinical final impression, which is typically at the end of the note.

🡪 If there is discordance between assertions at different points in the note, use the final impression/assertion, or the last assertion made in the note, to classify the document.

**b. Presence of X-ray/radiographic imaging report within the ED clinician note or discharge:**

Often, the imaging reports may be pasted into an ED clinician note, and this report contains an “Impression” made by the radiologist (ie. “Impression: no pneumonia”. In this case, location matters.

🡪 If the report exists above/outside of the relevant assessment/plan section, do not annotate these mentions of pneumonia; Scroll to the end of the ED clinician note to find the physician’s assessment/assertion.

🡪 If the report is placed within the assessment/plan section: If there is a positive assertion of pneumonia that uses clear pneumonia terms (“CXR positive for pneumonia) in the report and the clinician is using the report as evidence to support a diagnosis of pneumonia, then classify this as positive.

🡪 If a clinician raises the possibility of pneumonia by using clear pneumonia terms in their note, but then includes a chest imaging impression that has a *negative* assertion of pneumonia using clinical or radiographic terms (“CXR negative for infiltrate/consolidation; or CXR with no acute CP process; CXR normal) in the report, the clinician is using the report as evidence to refute a diagnosis of pneumonia, so classify this as negative.

**c. Broader diagnoses (sepsis/respiratory failure) that may have pneumonia as an etiology**: Patients -- especially sick ones -- sometimes have a clear problem such as sepsis or respiratory failure but without a clear cause by the end of the ED encounter. Pneumonia can be an etiology of these problems. However, unless the clinician raises the possibility of pneumonia as a potential etiology, the diagnosis of sepsis or respiratory failure are not sufficiently specific for a pneumonia diagnosis, and should not be classified as positive or possible.

**d. Diagnostic uncertainty or ambiguity**: If the clinician seems to be inconsistent in his/her assessment, take the latest-appearing statement as their final clinical suspicion.

It is typically useful to identify *where* the clinician asserts the diagnosis and to scan for terms for pneumonia.	Ex: If the initial impression is possible pneumonia but the final impression is clearly not pneumonia (for example if the clinician ruled pneumonia after physical exam and CXR), the document-level classification is clearly not pneumonia.

#### **3.5 Classifying a document in ehOST**

Select any word in the entire document by highlighting or double-clicking on it. Select “Document_Classification”. Click on attributes, and select the final classification from pull-down menu.

To classify pneumonia, highlight/annotate the last word or few words of the document, right-click, and choose the classification from the choices:

- Positive
- Possible
- Negative
- Not MD Note: This marks that a document should have been excluded from annotation

### 4. **Examples**

| Document class | Relevant Text | Reference/principle |
| --- | --- | --- |
| Positive | “Final Diagnosis: Pneumonia” |  |
|  | “A/P: Pna” |  |
|  | “A/P: Hospital-acquired pneumonia” |  |
|  | “A/P: Pneumonia resolving since last visit” | “Resolving” should be considered a positive term (as opposed to “resolved”) |
| Possible | “MDM: Differential diagnosis includes pneumonia, CHF” | Pneumonia is mentioned as a possible diagnosis but never confirmed. |
|  | “MDM: Differential diagnosis includes pneumonia, CHF.  Final Diagnosis: CHF  ” | Pneumonia is mentioned as a possible diagnosis. Although a different diagnosis is later assigned, pneumonia is never explicitly documented as being negative. |
|  | “MDM: rule out pneumonia. A/P: Chest x-ray was positive for infiltrate” | Pneumonia is mentioned as “Possible” and there is no negative evidence to negate it. |
|  | “MDM: possible pneumonia.  Labs/studies:  D-dimer normal  …  CXR: no infiltrate  EKG: …  Assessment/Plan: admit and treat for CHF” | Pneumonia is mentioned as “Possible” in a Tier 2 section. There are negative radiographic findings, but they are in the Labs/studies rather than a relevant section so cannot be used to exclude pneumonia.  Additionally, although an alternative diagnosis is given (CHF), pneumonia is never explicitly negated. |
| Negative | “Imaging: There is an infiltrate which is suggestive of pneumonia.” | Imaging text cannot be used as confirmed evidence, it must be corroborated in a relevant section such as the A/P. |
|  | “MDM: rule out pneumonia.  Assessment/Plan: Reviewed chest x-ray shown below:  Chest X-Ray: rule out pneumonia  Imaging impression: No infiltrate  Will admit and treat for CHF” | Pneumonia is mentioned as “Possible”, but subsequent negative radiologic findings which are included in the A/P rule it out. |
|  | “MDM: rule out pneumonia.  Assessment/Plan: Pneumonia is ruled out. Admit for CHF” | Pneumonia is listed as a possible diagnosis in a “Tier 1” section but is later explicitly negated in a “Tier 2” section. |
|  | “A/P: Chest x-ray was positive for infiltrate” | There are positive radiologic findings in the A/P but no clinical diagnosis or terms referring to pneumonia. |
|  | “A/P: Pneumonia is resolved” | “Resolved” is a negative term |

III. Radiology Report Annotation Guidelines

### **1. Background**

Thank you for helping to annotate for this project! Our goal is to develop a natural language processing tool that can classify chest imaging radiology documents on whether they contain a positive assertion of the radiographic diagnosis of pneumonia.

**The question we are asking is:**

***“Does the radiologist states/asserts that the image contains findings that could be consistent with a diagnosis of pneumonia in the appropriate clinical setting?”***

There are 2 imaging reports that we are targeting:

4. **Chest X-ray**: single 2-dimensional image that whitens against dense/”radio-opaque” matter (ie bones, fluid inc infiltrate) and darkens
5. **Chest CT (computed tomography)**: multiple X-rays rom 1000’s of angles, reconstituted as a 3-dimensional image that is represented as multiple “cuts” through which the viewer scrolls for review.

### **2. General Guidelines**

#### **2.1 Definitions: Positive, possible, or negative**

To allow the capture of some degrees of certainty, but acknowledge that certainty is not a discrete characteristic, we have 3 options for classification: “Positive”, “Possible” and “Negative”. For the Moore project, the NLP will focus on distinguishing between Positive/Possible from Negative. So it is more important for annotators to consistently distinguish positive/possible from negative.

1. **POSITIVE:**
   a. The radiologist states with clarity that there is an abnormality using terms included in the vocabulary list.
   b. The radiologist does not offer any additional diagnoses that better explain the abnormality. (ie, atelectasis, heart failure).
   c. The radiologist asserts a diagnosis of pneumonia (ie, “BEAR!”)
   d. Terms of uncertainty/certainty that we will classify as positive = “probable; suggestive of; consistent with; suspicious for.”
2. **POSSIBLE:**
   a. The radiologist describes an abnormality, and raises pneumonia, inflammation or infection as a possible explanation
   b. The radiologist does not describe with certainty that the abnormality is due to pneumonia/inflammation/infection, but uses terms that are included in the vocabulary list and does not offer an alternative explanation.
   c. Terms of uncertainty/certainty that we will classify as possible = “versus, possible, less likely, cannot rule out, may represent, and/or, or rather than”
3. **NEGATIVE:**
   a. The chest X-ray is stated as normal - “no acute cardiopulmonary process,” or “no infiltrate/consolidation.” Note that “stable” is not necessarily normal – review the entire findings section for abnormalities.
   b. The chest X-ray is abnormal, but the abnormality is clearly attributed to a diagnosis other than pneumonia (“ie linear opacity secondary to atelectasis”, “interstitial opacities secondary to CHF).
   c. Previous abnormalities consistent with infiltrates are noted to have clearly resolved. However, if terms “improving/improved, resolving” raise possibility that the infiltrates are still there, that image should be classified as possible/positive (see below).
   d. Terms of uncertainty/certainty that we will classify as negative = “ruled out, not”.

#### **2.2 Hierarchy of evidence**

Certain terms may be used to support either a diagnosis of pneumonia or other conditions. To account for this, we’ve established a hierarchy of **“Tier 1”** evidence (which is always interpreted to refer to pneumonia) and “**Tier 2”** evidence (which requires checking for other potential diagnoses.

| Tier 1 | Pneumonia  Consolidation |
| --- | --- |
| Tier 2 | Infiltrate  Opacity |
| Possible alternative diagnoses | Atelectasis  Pulmonary edema  Pulmonary fibrosis  Interstitial lung disease |

#### **2.3 Summary of classification logic**

- First, check if there is a mention of pneumonia or consolidation (Tier 1). If there is and it is definitively stated to be positive or negative (e.g., “no evidence of pneumonia”), classify the document as **“Positive”** or **“Negative”** accordingly.
- Next, check for a mention of infiltrate or opacity (Tier 2). If there is a negated mention or no mention, classify the document as **“Negative”**.
- If there is a positive mention of Tier 2 evidence, first check whether it is linked to another diagnosis. If it is not, classify the document as **“Positive”**. If it is, classify the document as **“Positive”**.

This logic is summarized in the diagram below.

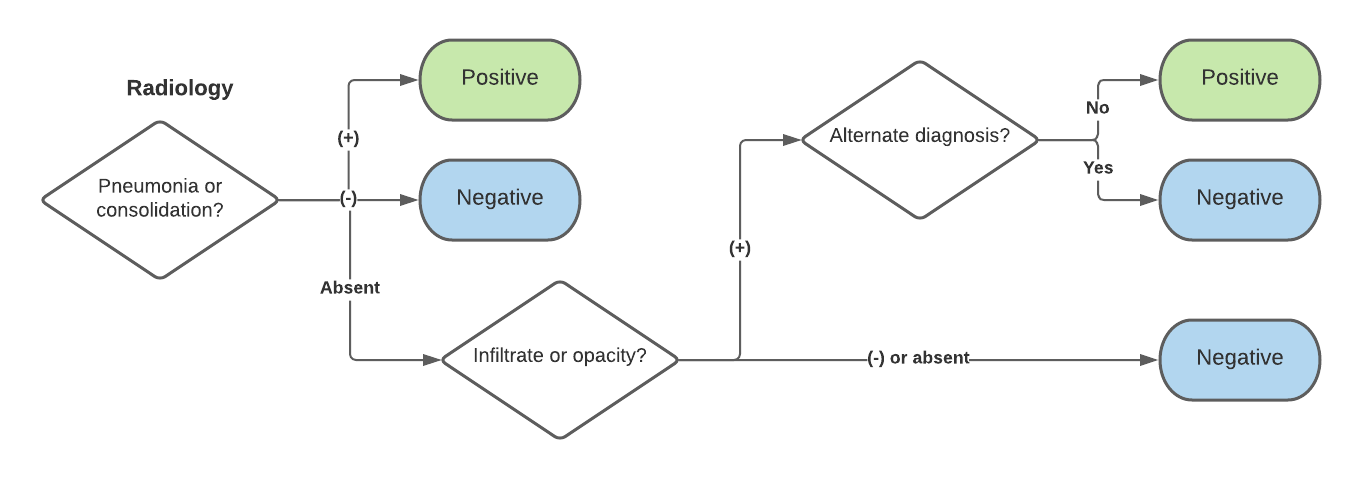

#### **2.4 General principles**

- **Location matters.**
  - The place in which the relevant information depends upon the note type.
  - For chest imaging reports, the sections are typically as follows:
    - STUDY//INDICATION//COMPARISON (any previous imaged they are comparing)// FINDINGS//IMPRESSION
  - The following sections usually have relevant information:
    - “FINDINGS” – detailed observations from the radiologist of the entire review
    - “IMPRESSION” – summary of main findings.
  - Relevant information will NOT typically be in “INDICATION” (an example: INDICATION: “pneumonia” means that the clinician ordered the image to evaluate for pneumonia, but does NOT mean that the radiologist found evidence to support this on the image.
- Terms used for radiographic assertions for pneumonia are similar to :
  - Pneumonia, pna, HCAP (“healthcare-associated pneumonia”)
  - Parapneumonic effusion = collection of fluid between lung and chest wall (the pleural space)
  - Empyema = collection of pus between lung and chest wall
  - If a clinician includes “infection/empyema” or “parapneumonic effusion” in their diagnosis, they are stating the patient has pneumonia.
  - For a (more) complete list, see **Section 3**.
- **Radiologists and certainty.** The tasks of the radiologist are to (1) describe what they see, (2) suggest potential explanations for what they see. They typically complete these tasks in succession. IE, (1) “I see a black furry thing in the forest” – (2) “I think it’s a bison. Or maybe a bear, less likely a moose.” Radiologists typically state (1) the findings with slightly more certainty, but typically do not assert (2) a clinical diagnosis with 100% certainty since you are supposed to combine the radiographic assertion with bedside clinical impression. There is variation in the degree to which the second task is felt to be warranted, and some radiologists are taught to avoid clinical diagnoses unless they have clinical input from the clinicians either directly or through the EHR. However if they are quite certain about it, they do sometimes call the diagnosis/etiology with certainty (“BEAR!”).
- **Comparison with previous images.**
  - For this task, we are **not** trying to classify documents based on whether the findings are new compared to a previous image. The task is NOT to identify a “new” infiltrate”.
  - If the radiologist indicates there has been an improvement: they may still indicate the infiltrate is persistent. Do NOT classify negative, unless radiologist clearly indicates that an infiltrate has **resolved (resolution, completely cleared**.) This includes strongly negative language such as “no acute cardiopulmonary process” or “no consolidation.” (If weak language around mentions of a previous infiltrate, and strong language that the image is negative, this should be classified as negative).

### **3. Specific terms**

Below is a (likely incomplete) list of terms which may be used to describe pneumonia in chest imaging reports. Remember to differentiate between Tier 1 and Tier 2 concepts as described above.

- Airspace disease
- Bronchiogram
- Bronchopneumonia
- Consolidation
- Consolidative density
- Haziness
- Increased lung/interstitial markings
- Infection
- Infectious process
- Infiltrate
- Infiltration
- Infiltrative
- Inflammation
- Inflammatory process
- Interstitial pneumonia
- Interstitial process
- Opacity
- Opacification
- Patchiness
- Pneumonia
- Pneumonic process
- Pneumonitis positive
- Reticulonodular pattern

IV. Discharge Summary Annotation Guidelines

Thank you for helping to annotate for this project! Our goal is to develop a natural language processing tool that can classify clinician-written documents on whether they contain a positive assertion of the diagnosis of pneumonia.

### **1. Background**

The document type we are targeting is the **Discharge summary**. This is a clinical document written by physicians at the end of a hospitalization, by physicians, nurse practitioners, or physician assistants who are responsible for asserting the final/ultimate diagnoses of pneumonia during a hospitalization – which were either present on admission (“community-acquired”) or developed during the hospitalization (“hospital-acquired”).

Other document types that are in the pipeline that using similar ontology include:

6. ED clinician Note: report written by emergency department clinician documenting an ED visit
7. Radiography report: report written by radiologist reviewing images: separate NLP with separate but slightly overlapping ontology.

Other document types that may eventually be used:

8. Admission History & Physical (written by admitting hospital care team)
9. Progress note (daily notes written by hospital care teams or consultants)
10. Consultation Report (initial note written by consulting service at the time of consultation)

**The question we are asking is:**

1. ***“Does the clinician/author of the note state/assert that the patient has/had pneumonia as a final diagnosis upon hospital discharge?***
2. ***If so:***
   a. ***Was this present at the time of initial hospital admission (“community-acquired pneumonia)”***
   b. ***Did this develop during the hospitalization? (“hospital-acquired pneumonia”)***

We are not asking:

- “Does the patient actually have evidence for pneumonia?” / “Do *you* think the patient actually had pneumonia?”/“Did the case meet a case definition for pneumonia” (ie, CDC/NHSN criteria)?
- “Does the patient have a history of pneumonia?”
- “Did the provider *treat* the patient as though the patient had pneumonia (ie mention antibiotics)”?
- “Did the provider *mean to say* that the patient had pneumonia” (ie, sepsis, respiratory source)?

The task of this classifier is to identify whether the clinician raise pneumonia as a diagnosis. We are looking for positive or possible assertions of a pneumonia diagnosis. **The task is NOT to accurately identify clearly negative assertions. (Thus the “negative” category means lack of assertion of a positive or possible pneumonia diagnosis, rather than an assertion that pneumonia was ruled out or excluded.)**

### **2. General Guidelines**

#### **2.1 Definitions: Positive, possible, or negative; hospital-acquired or community-acquired**

4. **POSITIVE:** Clinician states with certainty of the diagnosis of pneumonia as a final diagnosis of the hospitalization.
5. **POSSIBLE:** Possible pneumonia– clinician states the dx of pneumonia is possible or probable but uses terms of uncertainty.
6. **NEGATIVE:** Clinician indicates does not list pneumonia as a possible diagnosis or mentions that pneumonia was specifically excluded. Note: in this case, we are treating absence of evidence as evidence of absence… ie, “negative” just means a lack of positive/possible classification. (see below in the “inconsistencies/ambiguities” section for more details).

If a document is classified as “POSITIVE” or “POSSIBLE”, a secondary variable to extract from the document is whether it is hospital- or community-acquired. In eHOST, this is marked as an attribute for the document classification.

7. **COMMUNITY-ACQUIRED:** The clinician states that pneumonia was present at the time of initial hospital admission.
   a. **Note:** it is possible that pneumonia was present, but the diagnosis of pneumonia was not *made* at the time of admission by the ED physician (“missed” by the ED physician). This should still be classified as community-acquired.
   b. If neither community nor hospital-acquired are specified, the default assumption should be “community-acquired” and you don’t need to mark anything
8. **HOSPITAL-ACQUIRED:** if a document is classified as “POSITIVE” or “POSSIBLE” and the document also states that pneumonia developed during the hospitalization. A clear assertion of hospital-acquired pneumonia is made (terms= ventilator-associated, hospital-acquired, developed in hospital)
9. **BOTH**: Clinician states that the patient had pneumonia present on admission, then also developed another case of pneumonia during the hospitalization. This is rare, but can occur with long hospital stays, severe pneumonia that then develops a super-infection (ie, COVID complicated by ventilator-associated pneumonia).

#### **2.2 Summary of classification logic**

- A document is classified as **“Positive”** if pneumonia is definitively asserted as being a final diagnosis of the hospitalization within a relevant section such as “Diagnoses” or “Hospital Course”
- A document is classified as **“Possible”** if pneumonia is listed as a possible diagnosis of the hospitalization within a relevant section, but there is some uncertainty expressed. For example, “Final diagnoses: **Possible** **pneumonia**”
- A document is classified as **“Negative”** if:
  - pneumonia is not listed as a possible diagnosis within a relevant section; or
  - if pneumonia is explicitly negated (e.g., “**No** **pneumonia**”
  - ***Note***: This includes situations where a patient was initially assigned a diagnosis of pneumonia earlier in the hospitalization or ED visit but by the time of discharge it was ultimately changed (e.g., “Initially *thought to hav*e **pneumonia** but that **was ruled out/CT revealed pulmonary embolism**.”).
  - This also includes documents where pneumonia is asserted in a non-relevant section, such as the HPI, but not in a relevant section
- If a document is classified as **“Positive”** or **“Possible”** and it is specified to be **“Hospital-acquired”** or both community- and hospital-acquired, mark the “hospital-acquired” attribute accordingly

The logic is shown visually below. See **Section 3** for detailed terms and sections.

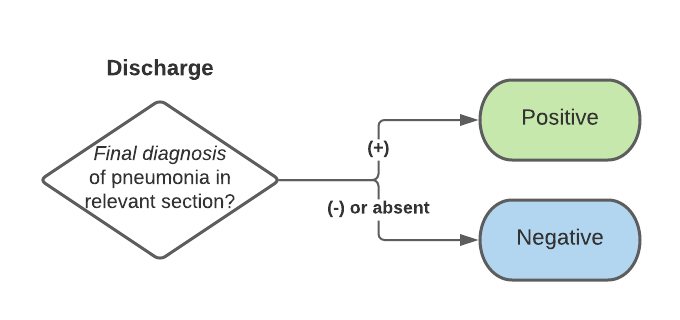

#### **2.3 Suggested workflow**

4. Find all relevant sections (see Table 1 for details).
5. Within these sections, search for clinical pneumonia terms.
6. If clinical pneumonia is mentioned in any section and not clearly excluded by a subsequent section, classify positive or possible accordingly.

#### **2.4 General principles**

**General Principles:**

- Use clinician’s narrative (assertion/suspicion within an assessment/plan), rather than objective information (evidence).
- Location matters: the place in which the relevant information depends upon the note type.
- Use what clinicians say, not what you think they mean. Ie, the task is to classify the clinicians’ statement/assertions of the diagnosis, but avoid inferring the diagnosis from their treatment decisions (ie, order CXR, treat with antibiotics).
- We are not trying to distinguish between new and existing cases of pneumonia. So a diagnosis of pneumonia acquired in a previous hospitalization or prior to admission could still be considered “Positive” unless it is asserted that it is resolved.
- Specific texts NOT to use when classifying:
  - Do not annotate chief complaint or reported symptoms
  - Do not annotate History of Present Illness.
  - Do not annotate assessment/plan, unless it is clearly located within the hospital course or the discharge diagnosis sections.
  - Do not annotate chest imaging report results
  - Do not annotate physical exam findings
  - Do not annotate vitals/labs.
  - Do not annotate medication lists that are not in the plan (ie the medications the patient was taking prior to the visit)
  - Do not annotate patient instructions/hypothetical (return to ER if…)

### **3. Specific terms and sections**

#### **3.1 Terms used for pneumonia**

| Clinical terms of pneumonia | Pneumonia and misspellings  HCAP  PNA  Bkp  bronchopneumonia  cap-  legionellosis  Parapneumonic effusion*  Empyema*  Pneumonia protocol |
| --- | --- |
| Radiographic terms of pneumonia  *These are more often used by radiologist, are non-specific to pneumonia; are positive for chest imaging, but cannot be used for DC summary. | Opacity  Infiltrate  Consolidation  Pneumonitis |
| Terms that may imply pneumonia but should **NOT** be used as specific pneumonia terms | Atypical infection  Respiratory infection  LRTI (lower RT infection)  Lung infection  Sepsis, lung source  Community-acquired |

***Parapneumonic effusion**: collection of fluid (effusion) between lung and chest wall (the pleural space) that is associated with a suspected pneumonia (parapneumonic).

***Empyema**: collection of pus between lung and chest wall.

If a clinician includes “infection/empyema” or “parapneumonic effusion” in their diagnosis, they are stating the patient has pneumonia.

#### **3.2 Linguistic modifiers**

Linguistic modifiers are phrases that refer to a mention of pneumonia and express uncertainty, negation, or temporality. When reviewing a mention of pneumonia, you should review for any of these terms to see if the mention of pneumonia should be excluded.

**Table 1. Linguistic modifiers for pneumonia**

| **Modifier category** | **Examples** |
| --- | --- |
| Positive existence | positive for  resolving  Improving  Consistent with |
| Possible existence | differential diagnosis  possible  Pneumonia vs *X*  X/pneumonia  rule out  Probable pneumonia  suspicious for  more likely pneumonia  concern for infection/empyema  Likely*  Suspicious for*  Suspect* |
| Negative existence | ruled out  no signs of  Resolved (prior to hospitalization – ie, did not occur during the hospitalization)  risk of pneumonia is low  is not likely  less likely  doubt  Excluded  Does not clinically correlate  Low suspicion for |
| Historical (annotate as “Negative”) | recent  past medical history of  resolved |

*Note: These terms express some remaining uncertainty but a likelihood that the diagnosis is pneumonia. They are classified as positive terms in the ED note, but only possible in the DC summary. This is because the DC summary is written at the end of the hospitalization, where overall certainty about the diagnosis is increased compared to the ED . ED clinical notes use more terms of uncertainty in general for this reason. If these terms are still used by clinicians authoring a DC summary, it has a greater meaning that the clinician was still not completely convinced that the diagnosis of pneumonia was correct.

#### **3.3 Relevant sections**

The *standard* organization of a Discharge summary is indicated here: <https://www.intel.com/content/dam/www/public/us/en/documents/white-papers/standardized-hospital-discharge-summaries-white-paper.pdf>

Although not all authors follow the standard organization, diagnoses are listed in the following sections:

- The **Diagnosis** section: “Final diagnoses”, “Principal diagnosis”, or “discharge diagnosis”
  - If pneumonia is present in “admitting diagnosis” but NOT present in the “final/discharge”, this may indicate that the author of the DC summary disagreed with the initial diagnosis. Look to the hospital course section. If the diagnosis was changed from pneumonia to another diagnosis, then this should be negative (no final diagnosis of pneumonia)
- The “**Hospital course**” section: the narrative of what happened during the patient’s hospital stay.
  - Note: the hospital course section can sometimes be named using a different term, such as “summary”. If it clearly contains the narrative of the hospital course (not the previous hospitalizations, medical history, or HPI), then look for assertions of pneumonia diagnosis within this section.
- NOT The H&P/HPI/ “history”/”brief HPI” portion: this will reflect the initial diagnosis, not the final diagnosis at discharge, so should be avoided. This also applies to “A/P/MDM” sections, unless they are clearly organized within the Hospital Course section.

Note that we are looking for an assertion of pneumonia within either of these sections. While the list of final diagnoses are often easier to identify and seem more certain, and are often the single part used for diagnosis coding, the assertion does not have to be present in both sections or the diagnosis section to be positive.

### **4. Setting a document classification**

#### **4.1 How to classify a document**

- Select the last word of the entire document by highlighting or double-clicking on it. Select “Document_Classification”. Click on attributes, and select the final classification from pull-down menu.
- To classify pneumonia, highlight/annotate the last word or few words of the document, right-click, and choose the classification from the choices:
- DOCUMENT_POSITIVE
- DOCUMENT_POSSIBLE
- DOCUMENT_NEGATIVE

#### **4.2 Setting “community”- vs “hospital”-acquired (Positive/Possible ocuments only)**

- For positive and possible documents, if the pneumonia was community-acquired/present on admission, then you do not need to classify anything further. However, if the pneumonia was specified hospital acquired, please indicate by changing an attribute to the document classification:
  - Select the highlighted span of text where you created an annotation
  - Select the magnifying glass in the lower right corner. This will open the “Attributes Editor”
- You should see an attribute called “location_acquired”. The default will be “community”. If needed, change it to “hospital” or “both”.

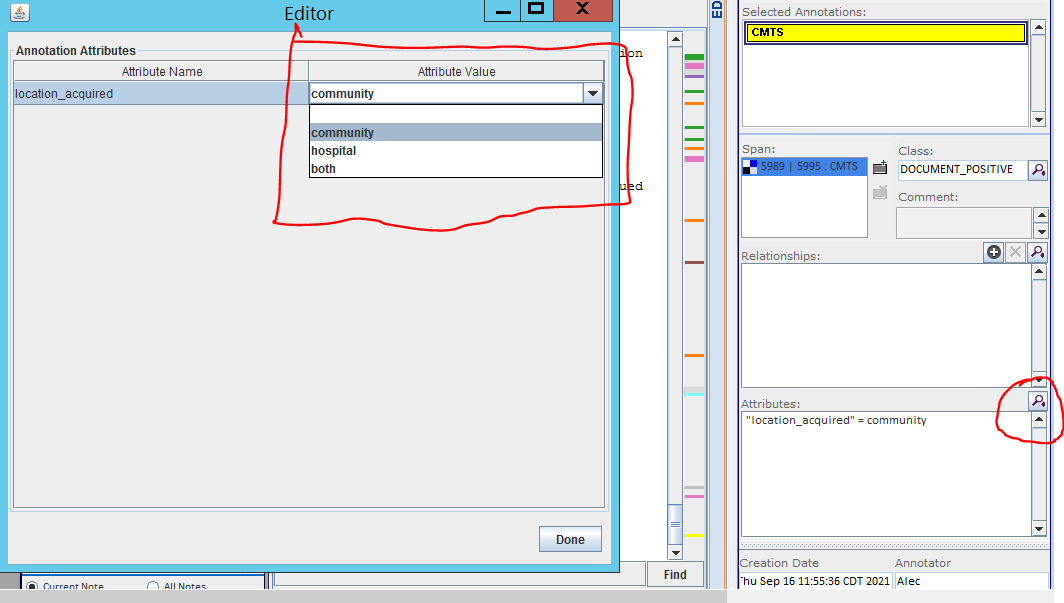

- If the note is clearly not a physician discharge summary (ex, it is an addendum showing XR result, discharge instructions, medication reconciliations, nursing note, or lab results etc), classify as “DOCUMENT_BAD_NOTE” or “DOCUMENT_NEGATIVE”. The NLP will ultimately need to process some documents like this, so we will consider such annotations “Negative”.
